# Healthcare workers in elderly care: a source of silent SARS-CoV-2 transmission?

**DOI:** 10.1101/2020.09.07.20178731

**Authors:** Mirjam Dautzenberg, Andrea Eikelenboom-Boskamp, Miranda Drabbe, Jacqueline Janssen, Ewoud de Jong, Eefke Weesendorp, Marion Koopmans, Andreas Voss

## Abstract

**Importance:** Healthcare workers (HCWs), including those with mild symptoms, may be an important source of COVID-19 within elderly care.

**Objective:** To gain insight into the spread of SARS-CoV-2 among HCWs working in elderly care settings.

**Design:** Cross-sectional study among HCWs working in elderly care in the South-East of the Netherlands, testing for SARS-CoV-2, between March 31 and April 17, 2020.

**Setting:** HCWs working in geriatric rehabilitation, somatic and psychogeriatric wards or small-scale living groups and district nursing, with a total of 5245 HCWs within 4 organisations.

**Participants:** 621 HCWs with mild respiratory symptoms.

**Main Outcomes:** Number of HCWs testing positive for SARS-CoV-2 in pharyngeal swabs, using realtime reverse-transcriptase PCR targeting the SARS-CoV-2 E-gene, N-gene, and RdRP. HCWs filled out a survey to collect information on symptoms and possible sources of infection.

**Results:** 133/615 (21.6%) HCWs tested positive for SARS-CoV-2, ranging from 15.6 to 44.4% per elderly care organisation, and from 0 to 64.3% per separate location of the organizations, respectively. 74.6% of tested HCWs were nursing staff, 1.7% elderly care physicians, 20.3% other HCWs with patient contact and 3.4% HCWs without patient contact. In the univariate analysis, fever, runny or stuffy nose, anosmia, general malaise, myalgia, headache and ocular pain were associated with SARS-CoV-2 positivity, while gastro-intestinal symptoms and respiratory symptoms, other than runny or stuffy nose were not. Risk factors for SARS-CoV-2 positivity were contact with patients or colleagues with suspected or proven COVID-19. Whole genome sequencing of 22 samples in 2 facilities strongly suggests spread within facilities.

**Conclusions and Relevance:** We found a high SARS-CoV-2 prevalence among HCWs in nursing homes and district nursing, supporting the hypothesis of undetected spread within elderly care facilities. Structural testing of elderly care HCWs, including track and trace of contacts, should be performed to control this spread, even when only mild symptoms are present.

## INTRODUCTION

On February 27, 2020, the first COVID-19 patient was detected in the Netherlands.^1^ On March 31, there were 12,595 Dutch patients known to be SARS-CoV-2 positive.^2^ As of early March, healthcare workers (HCWs) in acute-care settings, including those with mild symptoms, were widely tested, whereas public health services followed different testing strategies for other HCWs. At that time, the public health testing strategy included testing the first two residents with symptoms suggesting COVID-19 within a cohort in an elderly care facility and in case of positive results, precautions for the entire ward were taken. Testing of HCWs was not routinely performed. On March 19, a national policy was launched to ban all visitors to elderly care facilities. Our facilities implemented the ban with exception of end-of life-situations and in case of serious behavioural problems. At about that time all HCWs were asked to wear a medical mask in case of mild symptoms. Also, patients with respiratory symptoms without contact with COVID-positive patients, or travel to endemic region were considered to be at risk of having COVID-19. As of April 6, the national public health strategy was changed, to include testing of HCWs in non-acute settings in case of fever and/or respiratory complaints. Preceding this policy change, we tested HCWs in our regional elderly care facilities and district nursing, to gain insight into the spread of SARS-CoV-2 among HCWs within elderly care, including symptoms and risk factors for acquisition of SARS-CoV-2.

## METHODS

### Study design

A cross-sectional study was performed among HCWs working in elderly care in the South-East of the Netherlands, testing for SARS-CoV-2. In total 621 (11.8%) HCWs were tested spread over four organizations with a total of 5245 HCWs; 536 HCWs working in geriatric rehabilitation, somatic and psychogeriatric wards or small-scale living groups spread over 46 locations, and 85 HCWs working in district nursing in two out of four organizations. Written informed consent was obtained from all HCWs. Analyses were performed on de-identified data.

### Study population

HCWs with mild respiratory symptoms (not included in the case definition of COVID-19 at that time) were voluntarily tested between March 31 and April 17, 2020. HCWs were selected based on necessity for continuity of care or concerns with HCW’s health status. While the study was primarily intended to only include HCWs with mild symptoms, not included in the case definition of COVID-19 at that time, nursing homes used the opportunity to finally get their HCWs tested, as the public health services policy at this time only tested the first two cases per unit. Consequently, the included population became a mixture of HCWs with mild to moderate symptoms.

### Procedures

#### Survey

At the moment of testing, HCWs filled out a survey to collect information on symptoms and possible sources of infection. Information was collected on general non-respiratory symptoms (general malaise, anosmia, fever, myalgia, ocular pain, headache, chest pain), respiratory symptoms (runny or stuffy nose, coughing, dyspnoea, sore throat) and gastro-intestinal symptoms (abdominal pain, vomiting, diarrhoea or loose stools), possible sources of infection (attendance to event >50 people, travel abroad, contact with persons suspected or positive for SARS-CoV-2 infection (patients, colleagues, household members or others)), and date of start of symptoms.

#### PCR

Pharyngeal swabs were collected by dedicated personnel, and samples were sent to Wageningen Bioveterinary Research for real-time reverse-transcriptase PCR targeting the SARS-CoV-2 E-gene, N-gene, and RdRP. Extraction was performed on the KingFisher Flex (Thermofisher) with the ID Gene Mag Fast Extraction Kit (ID-Vet Genetics), with an input volume of 145 μl sample and 150 μl lysis buffer, and an output volume of 60 μl. The extraction was internally controlled (duplex PCR) using 5 μl green fluorescent protein (GFP)-RNA. Amplification and detection was performed on the QuantStudio5 (Applied Biosystems) with a cycling profile of 10 min at 52°C, 3 min at 95°C, 45 cycles of 15 sec at 95°C and 30 sec at 58°C. Extracted nucleic acids were amplified using TaqMan Fast Virus 1-step Master Mix (Applied Biosystems), and primer and probe mixture for the E gene as described previously (0.4 uM/primer, 0.2 uM probe).^3^ Analyses were performed using QuantStudio5 Design & Analysissoftware v 1.4.3 (threshold 0.1, and visual check of curves). In case of inconclusive PCR results, HCWs were retested.

#### WGS

Whole genome sequencing (WGS) was performed on a convenience sample of pharyngeal swabs, including samples from known COVID-19 nursing home residents at the corresponding locations,. Complete genome sequences were generated by SARS-CoV-2 specific, amplicon-based Nanopore sequencing, as previously described.^4^ Sequences were aligned and analysed against the background of a nationally representative set of genomes as described.^4^ Analyses were performed using a maximum likelihood tree.

### Statistical analysis

Continuous variables are expressed as medians and ranges. Categorical variables are expressed as counts and percentages. No formal sample size calculation was performed. Groups were compared using Pearson’s Chi-square test, Fisher’s exact test in case of expected counts <5, or Mann-Whitney-U test, and p-values <0.05 were considered statistically significant. Risk ratios were calculated to determine effect size. All analyses were performed with SPSS version 25 (IBM, Armonk, NY, USA).

## RESULTS

A total of 621 HCW were tested for SARS-CoV-2, of which six had inconclusive RT-PCR results and were excluded from analyses. Of the 615 remaining HCWs, 133 (21.6%) tested positive (2.5% of all HCWs from the 4 elderly care organisations). The positive HCWs were from all (n=4) elderly care organisations, and from 18 out of 46 (39.1%) locations, respectively. In case of incidental missing values, HCWs were still included, therefore denominators differ throughout the paper. Ten cases with major omissions in the survey were deleted completely from analyses.

Per location, a median of five HCWs were tested, ranging from 1 to 83. The percentage of HCWs infected with SARS-CoV-2 per elderly care organization ranged from 15.6 to 44.4%, and from 0 to 64.3% per separate location of the organizations, respectively.

444 (74.6%) of tested HCWs were nursing staff, 10 (1.7%) elderly care physicians, 121 (20.3%) other HCWs with patient contact (such as nutrition- and living assistants, cleaners) and 20 (3.4%) HCWs without patient contact (Table 1). Median age was 48.7 years, and 6.1% was male. The majority of tested HCWs experienced coughing (67.8%), runny or stuffy nose (66.6%), and general malaise (66.4%).

**Table 1.** Demographics and symptoms of HCWs tested for SARS-CoV-2, by PCR result.

|  | Positive (n=131) |  | Negative (n=474) |  |
| --- | --- | --- | --- | --- |
| **Demographics** |  |  |  |  |
| Male | 9 | 6.9% | 28<sup>a</sup> | 5.9% |
| Age (years) | 48.9 | 18.5-65.0 | 48.6<sup>a</sup> | 19.4-68.6 |
| **Profession<sup>b</sup>** |  |  |  |  |
| Medical doctor | 0 | 0.0% | 10 | 2.1% |
| Nurse | 94 | 72.9% | 350 | 75.1% |
| Other with patient contact | 35 | 27.1% | 86 | 18.4% |
| HCW without patient contact | 0 | 0.0% | 20 | 4.3% |

Results are given as n (%), or median (range). <sup>a</sup> Missing value for one HCW. <sup>b</sup> Missing values for 2 positive and 8 negative HCWs.

In univariate analysis, fever, runny or stuffy nose, anosmia, general malaise, myalgia, headache and ocular pain were associated with SARS-CoV-2 positivity (Table 2, p<0.05). Gastro-intestinal symptoms and respiratory symptoms with the exception of runny or stuffy nose were not associated with a positive SARS-CoV-2 test.

**Table 2.**
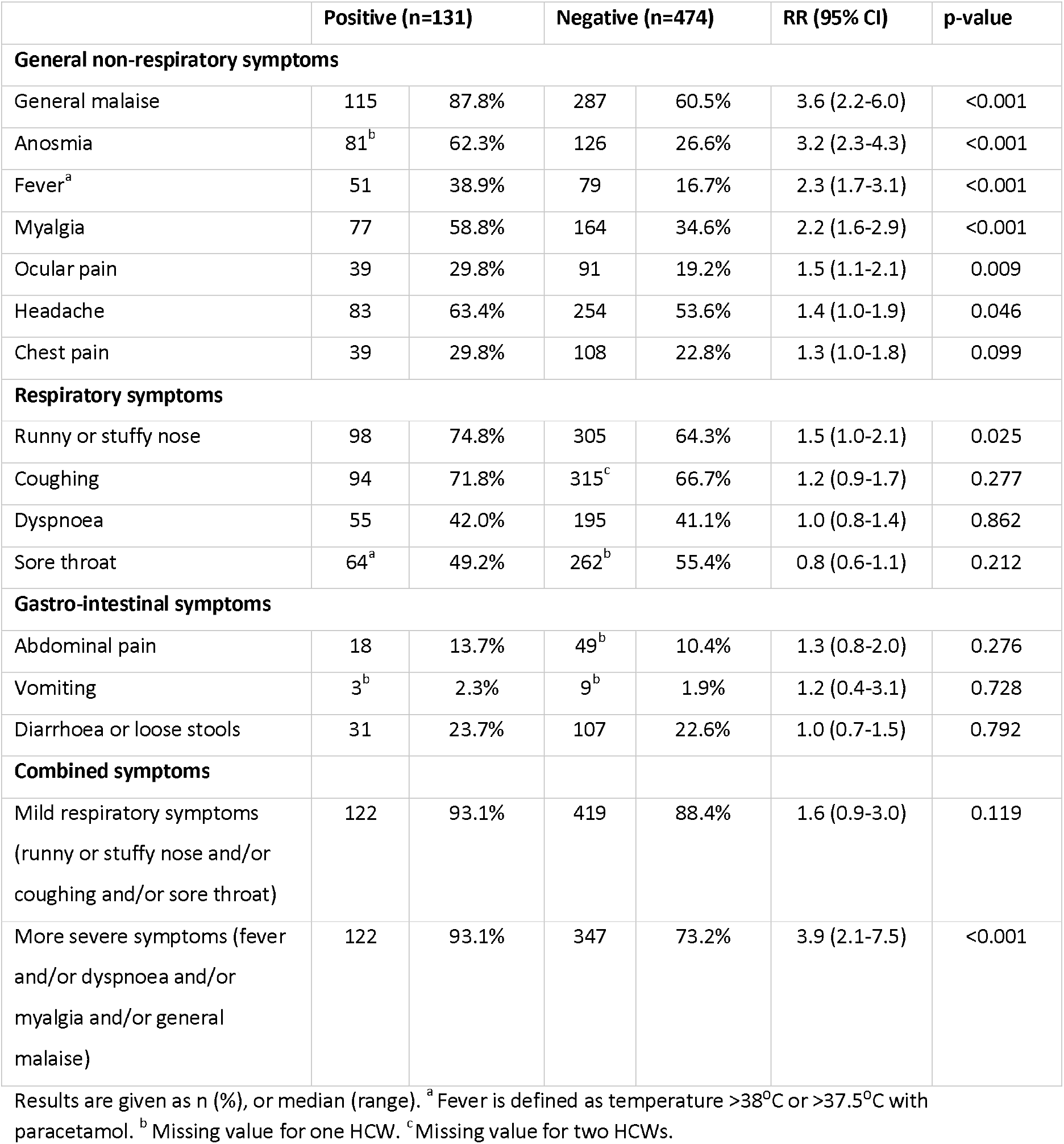
Symptoms of HCWs testing positive and negative for SARS-CoV-2.

SARS-CoV-2 positive HCWs without fever (n=80) presented more often with runny or stuffy nose than HCWs with fever (n=51) (83.8 vs 60.8%). They also report more often working with complaints (75.0 vs 50.0%). SARS-CoV-2 positive HCWs with fever more often presented with general malaise, myalgia and headache than HCW without fever (96.1 vs 82.5%; 72.5 vs 50%; and 74.5 vs 56.3% respectively, p<0.05).

In our population, attendance to events with more than 50 people, and travel abroad the last 14 days before start of symptoms, were not related to a positive SARS-CoV-2 PCR (Table 3). SARS-CoV-2 positive HCWs more often had contact with any person either proven (63.1 versus 37.7%) or suspected of (71.5 versus 48.1%) COVID-19 than those not infected. HCWs infected with SARS-CoV-2 significantly more often reported contact with patients or colleagues with suspected or proven COVID-19 than those not infected. No difference was seen in contact with proven or suspected household members or other contacts.

**Table 3.**
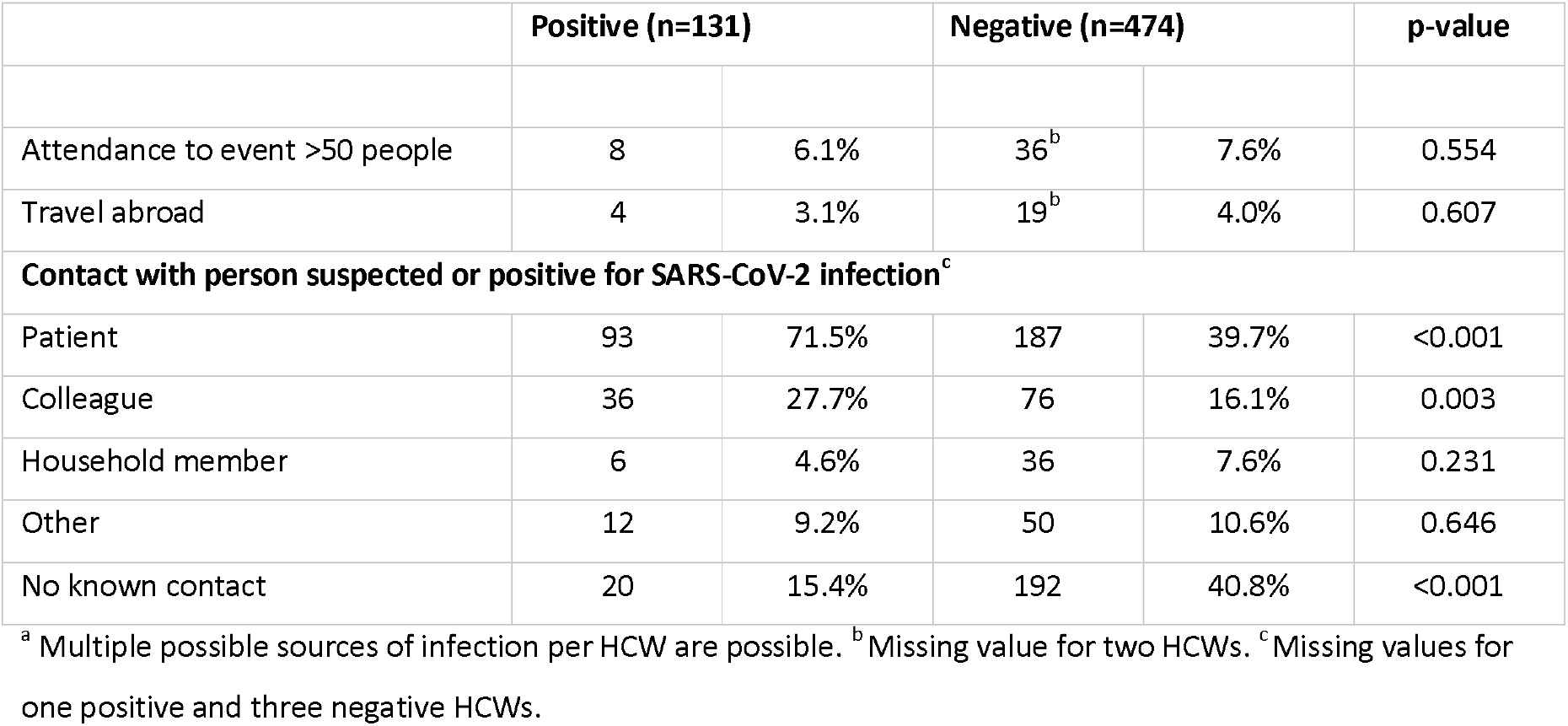
Possible sources of infection within 14 days before start of symptoms^a^

|  | Positive (n=131) |  | Negative (n=474) |  | p-value |
| --- | --- | --- | --- | --- | --- |
| Attendance to event >50 people | 8 | 6.1% | 36<sup>b</sup> | 7.6% | 0.554 |
| Travel abroad | 4 | 3.1% | 19<sup>b</sup> | 4.0% | 0.607 |
| **Contact with person suspected or positive for SARS-CoV-2 infection<sup>c</sup>** |  |  |  |  |  |
| Patient | 93 | 71.5% | 187 | 39.7% | <0.001 |
| Colleague | 36 | 27.7% | 76 | 16.1% | 0.003 |
| Household member | 6 | 4.6% | 36 | 7.6% | 0.231 |
| Other | 12 | 9.2% | 50 | 10.6% | 0.646 |
| No known contact | 20 | 15.4% | 192 | 40.8% | <0.001 |

<sup>a</sup> Multiple possible sources of infection per HCW are possible. <sup>b</sup> Missing value for two HCWs. <sup>c</sup> Missing values for one positive and three negative HCWs.

Median reported duration of symptoms before testing was 7 days (range 1-44 days) in SARS-CoV-2 positive HCW, and 11 (range 0-53) days in SARS-CoV-2 negative HCWs (p<0.001), and is depicted in Figure 1. About 1 out of 10 (11.8%) of the HCWs were no longer able to report the first day of symptoms. One of eight (12.5%) HCWs tested on the first reported day of symptoms, tested positive. For days 2-7, 8-14 and >14, 72/215 (33.5%); 31/141 (22.0%) and 17/169 (10.1%) HCWs tested positive, respectively. In 73 (13.7%) HCWs the reported duration of symptoms was > 21 days, of which seven tested positive. In total 391 (65.6%) HCWs report to have worked while symptomatic, with no difference between HCWs testing positive or negative.

**Figure 1.**
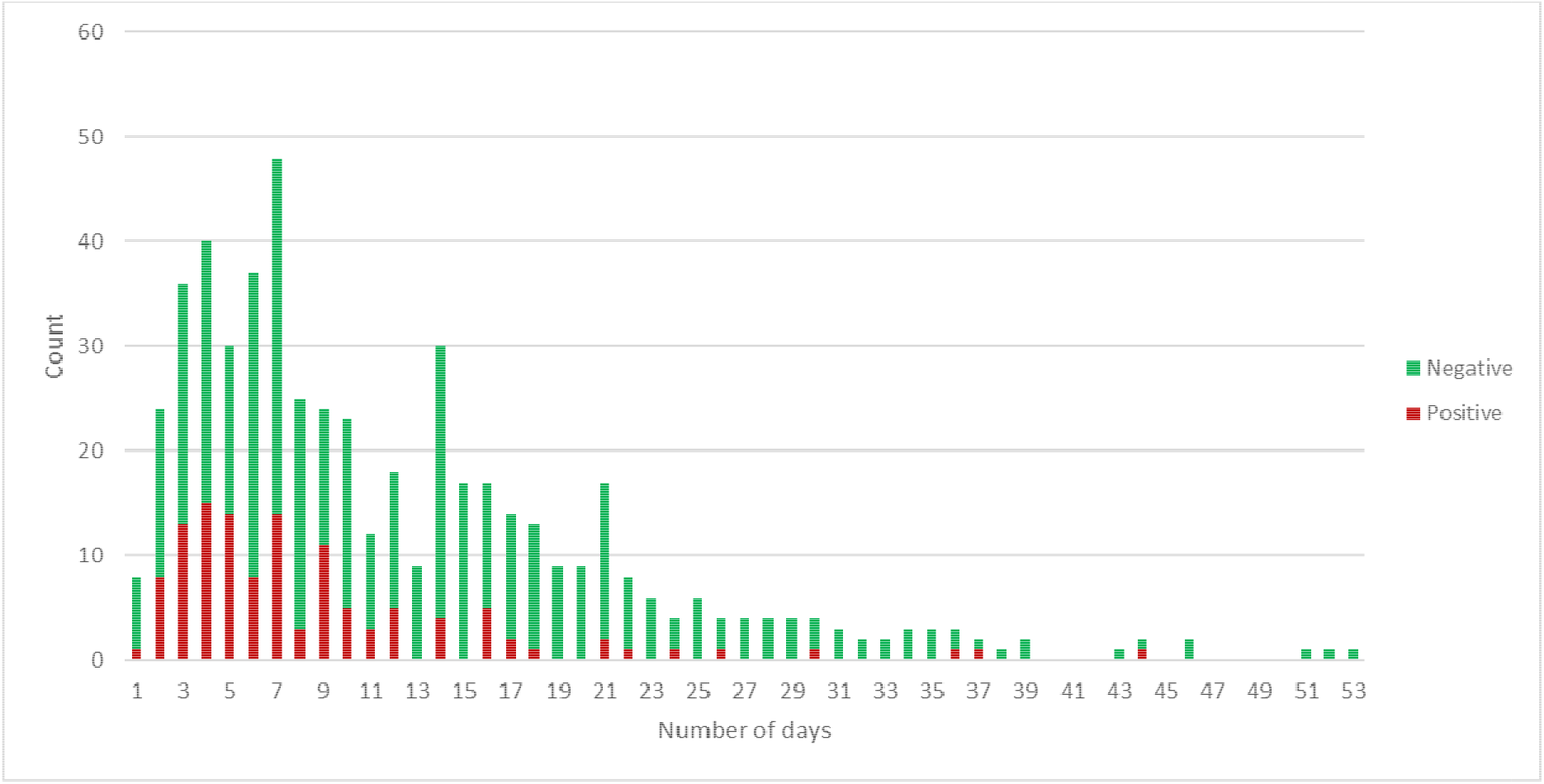
Reported duration of symptoms preceding testing of 533 HCWs in elderly care facilities.

### WGS

WGS was performed on 9 samples from one location, and 13 samples from another location. Two patients and seven HCWs from the first location cluster together, and five patients and seven HCWs from the second location cluster together, strongly suggesting spread within the nursing home. Only one patient in the second location had a unique strain, suggesting a separate introduction.

## DISCUSSION

In a sample of 621 HCWs with mild complaints working in 44 different nursing homes and district nursing, 21.6% tested positive for SARS-CoV-2. This high prevalence supports the hypothesis of undetected spread within elderly care facilities. Using WGS we documented the spread between patients and HCWs within two facilities with positive HCWs and patients.

We found a larger proportion of HCWs positive than the prevalence of 6% of COVID-19 amongst 86 symptomatic hospital HCWs in two Dutch hospitals^5^, and 9% of tested HCW in a university medical centre in our region^6^. There are a few possible explanations for this high prevalence. Our study was performed at a later time, when the prevalence and community spread of COVID-19 was higher in the Netherlands. Hospitals and elderly care facilities differ in the fact that in elderly care facilities there is a strong emphasis on living conditions and social interactions, and until closing of the facilities on March 19, introductions into the facilities could take place not only through HCWs, but also through visitors, and residents visiting places outside the facility, providing opportunity for repeated introduction of SARS-CoV-2 through individuals infected in the community.^5^ At the time of our study, nursing home clients with respiratory symptoms were only considered to be at risk of having COVID-19 when they had been in contact with a proven COVID-19-positive patients, or travelled to an endemic region. Mild symptoms such as anosmia or atypical symptoms such as diarrhoea were not recognized as symptoms for COVID-19. In addition, the use of personal protective equipment (PPE) such as masks in the nursing homes, was, at this time, limited to positive or suspected patients and even within that group not always adequate.

The majority of the tested HCWs (66%) report to have worked while symptomatic, which is comparable to 63% recently reported in Dutch hospitals.^5^ In addition, only as of mid-March, HCWs had to wear masks at all times in case of mild complaints. The large proportion of HCWs coming to work while symptomatic and the late introduction of masks for HCWs with mild symptoms, certainly contributed to preventable spread of SARS-CoV-2 in this setting.

Contact tracing is an important measure to detect and isolate infection sources and reduce transmission.^7^ Within the long-term care setting HCWs are less familiar with the measure, it is more difficult to perform, especially as low-threshold testing of patients was not available on the same scale as in hospitals.

Important risk factors that lead to the introduction of SARS-CoV-2 into the Netherlands, such as travel abroad and attendance of large-scale events (e.g. carnival), had no significant role (or no longer) in transmission of SARS-CoV-2 within our population of HCWs working in elderly care. At the time of our study COVID-19 was already widespread in the community.^4^ We did identify contact with patients or colleagues with suspected or proven COVID-19 as a risk factor for SARS-CoV-2 positivity, while no difference was seen in contact with proven or suspected household members or other contacts, suggesting that a significant proportion of infections in HCWs were acquired in the elderly care setting.

We identified fever, runny or stuffy nose, anosmia, general malaise, myalgia, headache and ocular pain to be associated with SARS-CoV-2 positivity, which is largely in line with symptoms identified in a recent Dutch study in hospital-based HCWs.^6^ Gastro-intestinal symptoms and respiratory symptoms with the exception of runny or stuffy nose were not associated with a positive SARS-CoV-2 test, and cannot be used in differentiating between a positive and a negative test within HCWs with complaints.

Fever was only present in 38.9% of HCWs with COVID-19, so other symptoms should be included to identify HCWs that should be tested. We used the definition of mild respiratory symptoms (runny or stuffy nose and/or coughing and/or sore throat) to identify HCWs that should use a medical mask during patient contact. 93.1% of SARS-CoV-2 positive HCWs met this definition, however, also 88.4% of SARS-CoV-2 negative HCWs, and this definition is therefore not discriminating between SARS-CoV-2 positivity and negativity. We used a definition of more severe symptoms (fever and/or dyspnoea and/or myalgia and/or general malaise) to identify HCWs that were banned from work, and should be tested. This latter definition was associated with a risk ratio of a positive test of 3.9 (95% confidence interval 2.1-7.5, p<0.001).

Seven HCWs report to have had symptoms for an extended period (>21 days) before they tested positive. This can resemble actual long-time positivity, however, initial complaints might have been unrelated to COVID-19, and only mild additional symptoms developed in a later stage.

There are several limitations to this study. First, as HCWs were not familiar with nasopharyngeal swabs, throat swabs were used for testing, which might be less sensitive, and may have let to under detection of SARS-CoV-2.^8,9^ Also, as a consequence of the cross-sectional nature of our study, timing of swabs was not optimal, probably leading to underdetection of HCWs testing too late in the course of their disease. More studies are needed to determine the prevalence of COVID-19 in this population, possibly based on serological testing.

In this study we showed a large previously undetected pool of COVID-19 within elderly care settings, namely the HCWs. Structural testing of elderly care HCWs, including track and trace of contacts, should take place to control this spread, even when only mild symptoms are present.

## Data Availability

Data is available from authors upon request.

## CONTRIBUTORS

AV initiated the study. AE-B collected the data. MD analysed the data and wrote the first version of the manuscript. All authors revised the manuscript and approved the final version.

## ACKNOWLEDGMENTS

The authors thank the ZZG Zorggroep and the HCWs from the other 3 organizations, the medical microbiology laboratory of Canisius Wilhelmina Hospital and Wageningen Bioveterinary Research laboratory for co-organizing the routing of testing. Also, we thank the HCWs from all 4 organizations who took the samples of the HCWs. Last but not least, we thank all HCWs who have been tested and filled in the registration form.

